# Characteristics and outcomes amongst of older subjects from Long-term Care admitted with Stroke to Hospital

**DOI:** 10.1101/2025.04.23.25326324

**Authors:** Joseph Harbison, Joan McCormack, Olga Brych, Amy Lynch, Margaret O’Connor, Peter Kelly, Ronan Collins, Tim Cassidy

**Affiliations:** Dept of Medical Gerontology, Trinity College Dublin, Ireland; Irish National Audit of Stroke Care, National Office of Clinical Audit, Dublin, Ireland; University Hospital Limerick, Ireland; Department of Neurology, University College Dublin, Ireland; National Clinical Programme for Stroke, Health Service Executive, Ireland; St Vincent’s University Hospital, Elm Park, Dublin. Ireland

**Author notes:** **Corresponding Author:** Prof. Joseph Harbison, Dept of Medical Gerontology, Trinity College Dublin Mercer’s Institute for Research into Ageing., St James’s Hospital Dublin 8, Ireland.

**Keywords:** Stroke, Nursing Home Care, Acute Hospital Care, COVID19, outcome

## Abstract

**Introduction:** Internationally about 3% of people ≥65 years live in Long Term Care (LTC). We examined the characteristics and outcomes of people admitted from LTC with stroke nationally and how this changed over the COVID19 pandemic.

**Methods:** Data from Irish National Audit of Stroke 2019-2023 were analysed by source of admission. An age, sex and subtype matched control group was derived from patients admitted from home. Pre-stroke and discharge modified Rankin Disability Scores (mRS) were analysed.

**Results:** Of 25451 admissions, 891 (3.5%) came from LTC and 22393 (88.0%) from home, 864 (4.6%) of 18805 ≥65 years came from LTC. Patient’s median ages were higher from LTC (84 vs. 74 years) and there were more women (58.4% vs 42.6% (p<0.001, Chi Sq)). Ischaemic strokes (IS) constituted 750 (84.2%) of LTC and 19106 (85.3%) of home admissions (p=0.34).

LTC admissions declined significantly during the pandemic 2019 3.74%, 2020: 3.07%, 2021: 3.19, 2022: (3.58%) and 2023: (3.98%) (p=0.045 Chi Sq).

A lower proportion of LTC admissions than controls were independent pre-stroke (mRS<3) (17.1% vs. 73.5%) (Figure 1). Mortality was significantly higher for LTC residents (21.2% vs 17.3%, p=0.03). LTC patients were admitted less frequently to stroke units (60.4% vs 70.7%, p<0.001) but were equally likely to be thrombolysed (LTC: 8.9%, Home: 9.6% p=0.74). Admission from nursing home was not independently associated with discharge mRS on linear regression.

**Conclusion.:** Strokes from LTC had worse outcomes than controls and were less likely to receive organized care. The proportion of strokes from LTC declined during the pandemic.

## Introduction

In the 2022 the National Census of Ireland 776315 of 5149139 (15.1%) of people were aged ≥65 years [1]. Of these 23509 (3.0%) were resident in long term care (LTC) [2]. This is similar to data from other areas of Europe and the United States on residence in Long term care institutions and Nursing homes [3]. Older, institutionalised people have a higher rate of comorbidity, including stroke risk factors and a past history of stroke than the general population [4]. Population based studies have shown that Diseases of the Circulatory system, including stroke, represent more than 15% of these admissions [5]. A recent data linkage study [6] from Wales found that 7.0% of 86,602 individuals had suffered a stroke in the 12 months prior to Nursing Home admission and the incidence of stroke in the 12 months after was 26.2 per 1000 person years. Accordingly, strokes in people resident in Long term care and Nursing homes can represent a significant proportion of admissions to stroke services internationally, but little research has been done to characterise their frequency of admission, the level of care received and their outcomes in comparison to other groups of patients.

There is some dispute as to the effectiveness of hospital care on people admitted from Nursing Homes and Long-term care. A systematic review found that admission of such patients through Emergency departments was associated with a high mortality rate and a high complication rate [6]. In contrast, other work from Switzerland shows that people admitted from Nursing homes for surgical interventions had similar outcomes to patients admitted from the community [7]. There is, however very little published data on outcomes for people admitted from long term care environments with acute stroke despite the population living in LTC being more vulnerable. Previous studies that organized stroke unit care improves outcome for all subgroups and subtypes of stroke evaluated, but strokes in people from Long-term care and Nursing Homes have not been specifically included [8].

For more than ten years, the Irish National Audit of Stroke (INAS) has been collecting data on patients admitted with stroke to Irish Hospitals with organized stroke services [9]. People admitted to hospital from Nursing Homes represent an important and potentially vulnerable group of patients so we performed an analysis of available data to examine the demographics and characteristics of people from Nursing homes experiencing stroke and to compare their outcomes with those living in non-institutional community settings. The period of time we studied included the duration of the COVID19 pandemic which substantially affected Nursing Homes internationally and changed admission patterns from them [10,11]. We studied how admission pattens for stroke from LTC changed over the period of the pandemic in comparison with the general population.

## Methods

Five-year data from INAS from January 1st 2019 to December 31^st^ 2023 were analysed by source of admission as part of a cross-sectional, cohort study. This is a dataset routinely collected by clinical stroke teams in all hospitals admitting at least 25 stroke patients per annum [9]. The data is collected, primary for quality improvement purposes, to a national online database where it is merged with routinely collected demographic, process, discharge and outcome data from the Hospital In-Patient Enquiry (HIPE) which also records data on domicile prior to admission, including home, long term care and a diffuse range of other sources including hostel, hotel, prison or other hospitals, which for our purposes were characterised as ‘other’. The definition of long-term care used by HIPE is ‘nursing home/convalescent home or other long stay accommodation’. In respect of older people, the great majority of subjects in this classification come from either public or private nursing home care. Data on Ischaemic stroke (IS) and primary Intracerebral haemorrhage (PICH) were collected and analysed but data on sub-arachnoid haemorrhage is not systematically collected by INAS because varying pathways of management across the country result in different HIPE numbers and classifications and were not included.

An age, sex and subtype matched, randomly selected, control group was then derived from patients admitted from home. Where available, pre-stroke and discharge modified Rankin Scores (mRS) were analysed to assess baseline level of disability and outcome. All hospitals recruiting patients to INAS have physicians trained in geriatric and internal medicine participating in the management of people with stroke and the practice of Comprehensive Geriatric Assessment [12] of patients is widely implemented [13] and is of benefit in managing older people with stroke [14]. All of these geriatricians would have experience in caring for frail, Nursing Home residents.

Data were analysed using proprietary statistical software (SPSS v29) and using Microsoft Excel. Comparisons for continuous / quantitative data were made using Student’s t tests and for proportional data using Pearson’s Chi Square statistics. As the dataset was originally designed for the purposes of quality improvement, a number of covariates that may be available in an observational study or register are not currently collected e.g. comorbidities, body weight, impairment score which limits the utility of regression analysis. Some data points, notably pre-stroke and discharge mRS were not collected on a minority of patients, this data is presented in the accompanying results tables. At the time of this study the audit did not collect impairment data in the form of the NIH Stroke Score (NIHSS) and MRS is used to determine stroke outcome. Complete mortality data is also collected separately by HIPE as a discharge outcome.

Data on three measures of stroke care and process were compared between groups.

i. Admission to Stroke Unit.
ii. Whether a swallow screen was performed at admission.
iii. Whether patients with ischaemic strokes received thrombolysis. Data on thrombectomy was only integrated into the INAS dataset in 2023 and numbers were too small for analyses.

Although all data were anonymised prior to analysis, ethical approval was obtained for the study from the joint University Hospital institutional ethics committee (ref 4556) to comply with National Office for Clinical Audit (NOCA) institutional practice for data use outside the context of standard outcome reporting. INAS is a statutorily funded organisation and no external sources of funding were used for this study. All data used is freely available to review on application to NOCA at https://www.noca.ie/about-noca/access-to-audit-data/.

Data on Nursing Home Occupancy were collected from data published by the Irish Government Department for Public expenditure, which subsidises Nursing Home costs for most residents [15], and from the representational group for Nursing Home operators, Nursing Homes Ireland [16].

## Results

In the period 2019 – 2023, there were 25451 stroke admissions, of whom 891 (3.5%) came from LTC and 22393 (88.0%) from home (Table 1). The remaining 2167 (8.5%) came from other locations. There were 18805 individuals ≥65 years (73.9%) in the total population, from whom 864 (4.6%) were admitted from LTC. In 2023 alone there were 229 people admitted with stroke from nursing homes from a reported 31901 private and public Nursing Home beds in Ireland [15] with a reported 92.3% occupancy [16]. Thus, the calculated rate of admission for stroke from Nursing Homes to hospitals with organised services was 7.8/1000 person years.

**Table 1:** Characteristics of patients coming from each source: Home, Long term care, and other*.

|  | Long Term Care | Home | p value | Other |
| --- | --- | --- | --- | --- |
| N (% Total) | 891 (3.5) | 22393 (88.0) | <0.0001 | 2167 (8.5) |
| Median Age in years (IQR) | 84 (78-89) | 74 (64-82) |  | 70 (58-78) |
| ≥65 years (% total) | 864 (4.6) | 16563 (74.0) | <0.0001 | 1377 (63.5) |
| Women (%) | 520 (58.4) | 9542 (42.6) | <0.0001 | 845 (39.0) |
| Ischaemic Stroke (%) | 750 (84.2) | 19106 (85.3) | 0.34 | 1913 (88.3) |
| AF known admission (%) | 260 (29.2) | 3884 (17.3) | <0.0001 | 366 (16.9) |
| Total AF diagnosed (%) | 340 (38.2) | 5993 (26.8) | <0.0001 | 605 (27.9) |
| Inpatient death (%) | 189 (21.2) | 2351(10.5) |  | 186 (8.6) |
| Admitted to Stroke Unit (%) | 538 (60.4) | 15586 (69.6) | <0.0001 | 1715 (79.1) |
| Thrombolysed (% ischaemic) | 72 (9.6) | 1893 (9.6) | 1.0 | 256 (13.4) |
| Swallow assessed (%) | 677 (74.9) | 15458 (69.0) | <0.0001 | 1698 (78.3) |
| Admission mRS available | 805 (90.3) | 21055 (94.0) | <0.0001 | 2048 (94.5) |
| Median pre-stroke mRS (IQR) | 4 (3-4) | 0 (0-2) |  | 0 (0-0) |
| Pe-stroke mRS <3 (%) | 138 (15.5) | 18151 (81.1) | <0.0001 | 1867 (86.2) |
| Discharge mRS available (IQR) | 828 (92.9) | 20960 (93.6) | 0.57 | 2042 (94.2) |
| Median discharge mRS (IQR) | 4 (4-5) | 2 (1-4) |  | 3 (2-4) |
| Discharge mRS <3 (%) | 40 (4.8%) | 11721 (55.9) | <0.0001 | 782 (38.3) |
| Both mRS scores (%) | 798 (89.6) | 20770 (92.7) | 0.0003 | 2011 (92.8) |

Other\* includes transfer of admitted or non-admitted patient from Hospital or Covid -19 facility or transfer from any Acute Hospital not specified, transfer from Non-Acute Hospital, transfer from hospice, transfer from psychiatric hospital/unit, new born, temporary place of residence (e.g. hotel), prison or other location.

People admitted from LTC were typically older than those admitted from home (Median age LTC: 84 years, home: 74 years) (table 1). A greater proportion of those admitted from LTC were women (58.4% vs 42.6% (p<0.001, Chi Sq)). Ischaemic strokes constituted 750 (84.2%) of cases admitted from LTC and 19106 (85.3%) of those admitted from home (p=0.34 Chi Sq). Patients admitted from LTC were less likely to be admitted to a stroke unit (60.4% vs 69.6%). However patients admitted to stroke units from LTC had a lower in-hospital mortality (91/538 (16.9%) vs. 98/353 (27.8%) p<0.0001 Chi Sq). LTC patients admitted to stroke units were also more likely to have their swallow screened (447/538 (83.1%) vs. 250/353 (70.8%) p<0.0001 Chi Sq). Median pre-stroke mRS in those admitted from LTC was 4, suggesting a moderate to severe level of disability.

Patients from LTC were more likely to have Atrial Fibrillation (AF) identified (38.2% vs. 25.8%) largely resulting from a higher rate of previously known AF at admission (29.2% vs 17.3%) reflecting their older age. Data on pre-stroke and post-stroke mRS were available in about 90% of patients but pre-stroke mRS data was significantly more complete for those admitted from home (table 1).

Admissions from LTC dropped significantly during the peak pandemic years of 2020 and 2021: 2019 3.74%, 2020: 3.07% (18.3% relative drop), 2021: 3.19% (14.1% relative drop), 2022: (3.58%) and 2023: (3.98%) (p=0.045 Chi Sq). The proportion of LTC who died as an inpatient dropped non-significantly in the years 2020 and 2021 (p=0.09 Chi Sq) (figure 1).

**Figure 1.**
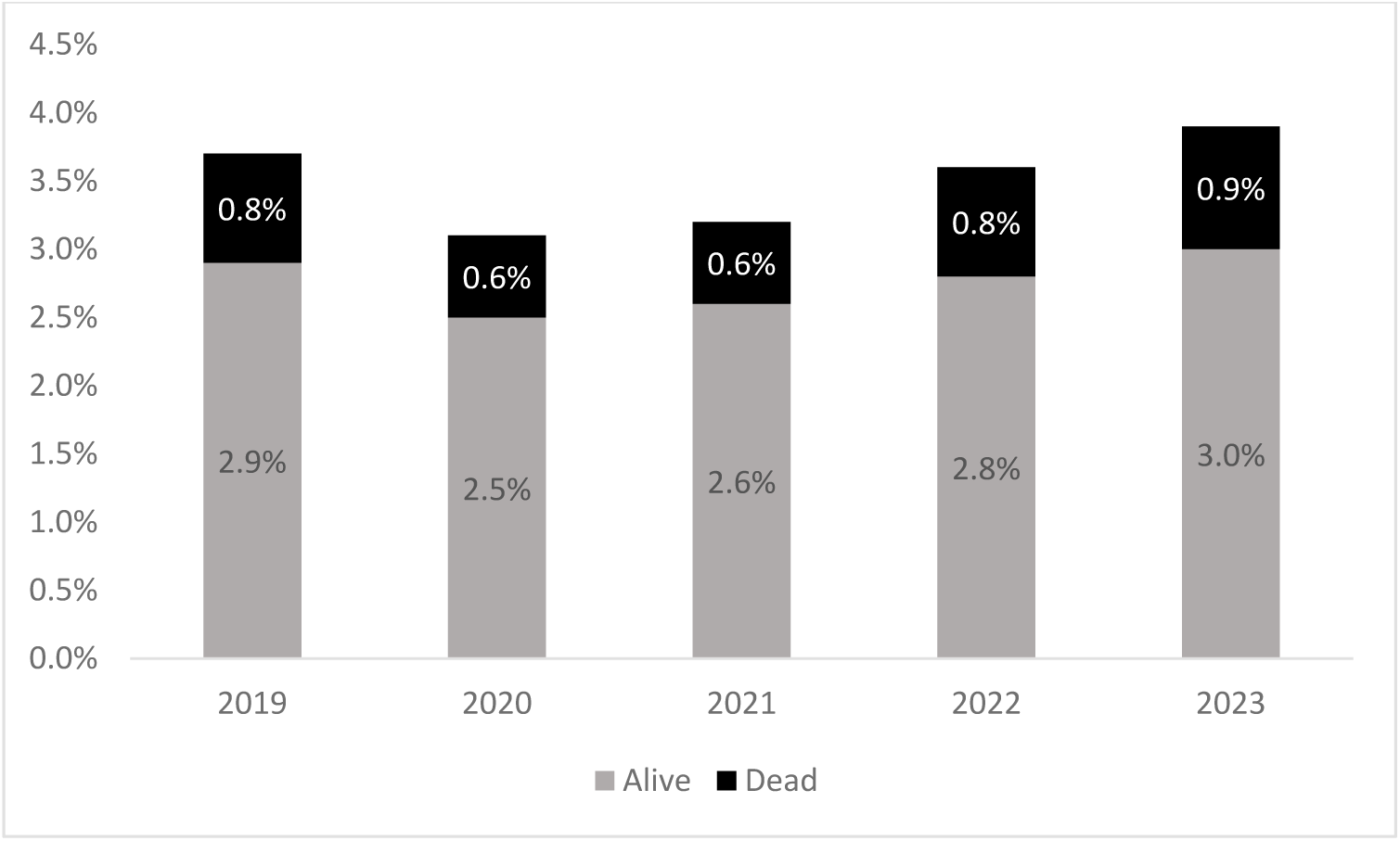
Changes in proportion of admissions (p=0.045 Chi Sq) and inpatient mortality (p=0.09 Chi Sq) of patients admitted from LTC with stroke over the period of the COVID19 pandemic.

When compared to controls (table 2), a lower proportion of those admitted from LTC were independent pre-stroke (mRS<3) (15.5% vs. 73.5%) (figure 2). In contrast, a higher proportion of those admitted from LTC recovered to their pre-stroke level of function (43.8%-31.9% p<0.0001) although this baseline level of function was typically much poorer (median mRS 4 vs. 1). Seventy eight of the 798 patients from LTC cohort (9.8%) with pre-stroke and discharge mRS data were reported as having a score of 0 or 1 pre stroke, suggesting full independence despite living in a care setting. Inpatient mortality was significantly higher (21.2% vs 17.3%, p=0.04 Chi Sq) in those admitted from LTC. Of the 135 patients from LTC that were independent (mRS<3) pre-stroke, 40 (29.6%) remained independent at discharge.

**Figure 2.**
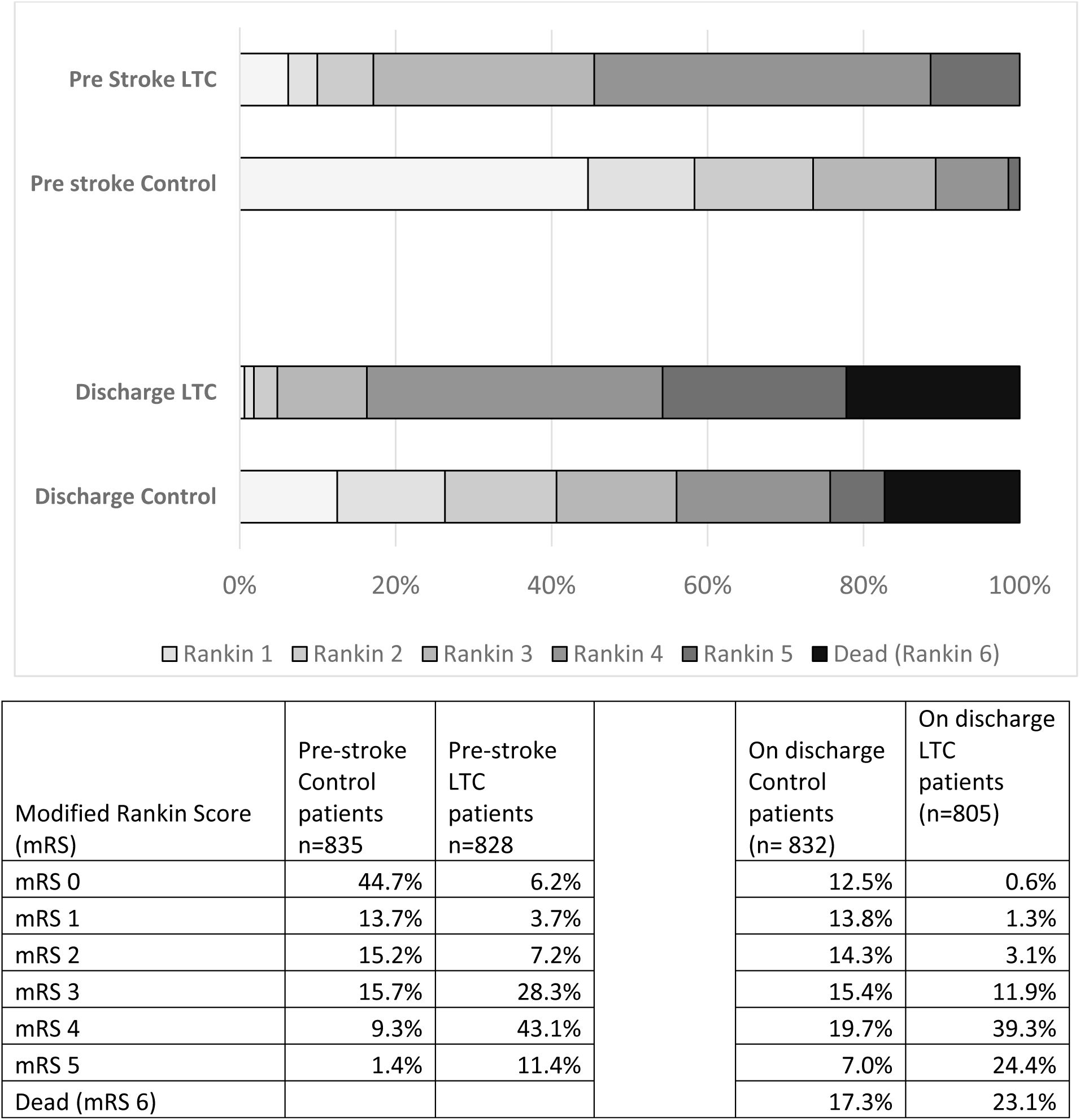
Comparison of pre-stroke and discharge modified Rankin scores between LTC cohort and age, sex and subtype matched control group admitted from home. Inpatient mortality (mRS 6) was significantly higher in the LTC cohort (p=0.02). Pre-stroke independence (mRS <3) was substantially higher in the control group 138 (p<0.0001).

**Table 2:** Comparison of outcomes between patients admitted from LTC and an age, sex and subtype (ischaemic or Haemorrhagic) matched cohort admitted from home.

|  | Long Term Care | Control | p value |
| --- | --- | --- | --- |
| N (% Total) | 891 | 891 |  |
| Median Age in years (IQR) | 84 (78-89) | 84 (78-89) |  |
| ≥65 years (% total) | 864 (4.6) | 864 (4.6) |  |
| Women (%) | 520 (58.4) | 520 (58.4) | <0.0001 |
| Ischaemic Stroke (%) | 750 (84.2) | 749 (84.1) | 1.0 |
| AF known admission (%) | 260 (29.2) | 242 (27.1) | 0.34 |
| Total AF diagnosed (%) | 340 (38.2) | 342 (38.4) | 1.0 |
| Inpatient death (%) | 189 (21.2) | 154 (17.3) | 0.04 |
| Admitted to Stroke Unit (%) | 538 (60.4) | 630 (70.7) | <0.0001 |
| Thrombolysed (% ischaemic) | 72 (9.6) | 67 (8.9) | 0.66 |
| Swallow assessed (%) | 677 (74.9) | 664 (72.3) | 0.48 |
| Admission mRS available | 805 (90.3) | 835 (93.7) |  |
| Median pre-stroke mRS (IQR) | 4 (3-4) | 1 (0-3) |  |
| Pe-stroke mRS <3 (% pre-stroke) | 138 (15.5) | 614 (73.5) | <0.0001 |
| Discharge mRS available (IQR) | 828 (92.9) | 832 (93.4) | 0.7 |
| Median discharge mRS (IQR) | 4 (4-5) | 3 (1-4) |  |
| Discharge mRS <3 (% Discharge) | 40 (4.8%) | 338 (40.5) | <0.0001 |
| Both mRS scores (%) | 798 (89.6) | 813 (91.2) | 0.20 |
| Returned to pre-Stroke mRS (% Both) | 350 (43.8) | 259 (31.9) | <0.0001 |

|  | B coeff | S.E. | O.R. | C.I. | p value |
| --- | --- | --- | --- | --- | --- |
| Admission Source | 0.104 | 0.060 | 1.11 | 0.99-1.25 | 0.083 |
| Discharge year | -0.024 | 0.008 | 0.98 | 0.96-0.99 | 0.002 |
| Age | 0.023 | 0.001 | 1.02 | 1.02-1.03 | <0.001 |
| Sex | 0.041 | 0.022 | 1.04 | 1.00-1.09 | 0.061 |
| Stroke Type | 1.393 | 0.030 | 4.03 | 3.79-4.28 | <0.001 |
| Pre-stroke mRS | 0.638 | 0.009 | 1.89 | 1.86-1.93 | <0.001 |
| Thrombolysed | -0.371 | 0.038 | 0.95 | 0.88-1.02 | <0.001 |
| Stroke Unit Care | -0.185 | 0.023 | 0.83 | 0.79-0.87 | <0.001 |
| Atrial Fibrillation | -0.210 | 0.025 | 0.81 | 0.77-0.85 | <0.001 |

|  | B coeff | S.E. | O.R. | C.I. | p value |
| --- | --- | --- | --- | --- | --- |
| Admission Source | 0.085 | 0.064 | 1.09 | 0.96-1.23 | 0.188 |
| Discharge year | -0.035 | 0.010 | 0.97 | 0.95-0.99 | <0.001 |
| Age | 0.038 | 0.002 | 1.04 | 1.03-1.04 | <0.001 |
| Sex | 0.021 | 0.028 | 1.02 | 0.96-1.08 | 0.449 |
| Stroke Type | 1.346 | 0.038 | 3.84 | 3.57-4.14 | <0.001 |
| Pre-stroke mRS | 0.604 | 0.011 | 1.83 | 1.79-1.87 | <0.001 |
| Thrombolysed | -0.405 | 0.049 | 0.67 | 0.61-0.73 | <0.001 |
| Stroke Unit Care | -0.223 | 0.030 | 0.80 | 0.75-0.85 | <0.001 |
| Atrial Fibrillation | -0.205 | 0.029 | 0.81 | 0.77-0.86 | <0.001 |

Patients admitted from LTC were less frequently admitted to stroke units than controls (60.4% vs 70.7%, p<0.0001 Chi Sq). In the control group, patients not admitted to Stroke Unit care had a higher inpatient mortality (93/630 (14.8%) vs. 61/261 (23.4%) p=0.002 Chi Sq) although lower than that found in LTC admissions. Patients admitted from LTC equally likely to be swallow screened as controls (74.9% vs 72.3% p=0.48 Chi Sq). Ischaemic strokes from LTC were equally likely to be thrombolysed than those from home (LTC: 9.6%, Home: 8.9% p=0.48). Prevalence of known AF and total AF diagnosed at discharge were not different but its notable that an additional 80 (9.0%) of the LTC patients had new AF diagnosed during admission, permitting effective secondary prevention be commenced.

On multivariate analysis (Table 3) using data from all patients admitted from home and LTC, admission from a Nursing Home was not significantly independently associated with discharge mRS (p=0.83)(Table 3). Highly significant associations (p<0.001) were however found with age, stroke type, pre-stroke mRS, year of admission if the individual had been thrombolysed, stroke unit admission and presence of AF.

On repeat linear regression (Table 3B) including only subjects ≥70 years again, admission source was again not independently associated with discharge outcome. In neither analysis was patient sex significantly associated with outcome.

## Discussion

The proportion of patients admitted with a stroke from nursing homes exceeded the proportion of the national population resident in LTC, even when considering only those ≥65 years of age. The decline found in stroke admission from Nursing Homes through the COVID19 pandemic will have slightly reduced the overall admission rate in this study. Nursing home residents were not significantly less likely to undergo thrombolysis or have their swallow safety checked than age and sex matched controls, but they were also less likely to be admitted to organised stroke unit care. Although the patients from long LTC were substantially more disabled at baseline than those admitted from home, inpatient mortality was only moderately higher and on multivariate analysis, admission from Nursing home was not itself a predictor of poorer outcome.

There are limitations to this form of study. INAS data is collected as a routine by hospital-based stroke teams but there is a higher proportion of missing data than would be seen in the context of a specific research database or register. This was particularly the case for missing mRS data both pre and post stroke and this was more noticeable in people who came from Nursing Homes than those who came from home. We suspect that it was easier to determine that a person was fully independent at home pre-stroke than to ascertain the level of disability of a person in Long term care. Another limitation is that INAS is hospital based and collected by stroke teams so only collects data from acute general hospitals with stroke services. It is conceivable that older patients in LTC who experienced massive stroke in LTC even at home, or those who have significant other comorbidities may not be referred into hospital as a decision by their families or family doctor. Similarly, people may choose not to refer in people with only minor impairments if they occur in the context of other comorbidity like dementia or frailty if their home or residential facility is remote to the hospital. A previous study in North Dublin in Ireland found that 9.5% of people suffering stroke did not present to hospital [17] but we feel that proportion is likely to be higher both in the age group under evaluation and in non-urban areas with less convenient access to hospitals.

We specifically did not analyse data from patients admitted from locations other than LTC facilities or their own home. The ‘other’ category is complex as it predominantly includes interhospital transfers e.g., after procedures, or transfer from hospitals with no stroke services. We have no record of where these patients may have originated from prior to their initial hospital admission thus effective analysis could not be conducted.

It is recognised that term care facilities are associated with a higher risk of stroke than the community for multiple reasons, but there is a paucity of epidemiological data as to stroke and stroke outcome in patients transferred to hospital from nursing homes or other care facilities. People admitted to LTC have more disability and comorbidity than those still resident in the community and the prevalence of previous stroke in Irish Nursing homes has been estimated to be around 17%-18%, meaning that disabling stroke is a common reason for LTC admission [18,19] but what proportion of stroke is acquired as a resident in LTC is unknown.

The decline in admission rate during 2020 and 2021 was associated with efforts to reduce movement in and out of Nursing Homes to prevent spread of COVID19 [20]. While the overall proportion of admissions from Long term care settings was small, the fall in proportion of admissions between 2020 and 2021 equated to about fifty individuals not admitted to hospital with stroke. This raises a question about non-transfer of patients from Nursing Homes to hospital following stroke in other circumstances. Clearly in many cases a decision may be made by a physician is association with the patient and their family, that such a transfer may not be appropriate given the individuals preferences and underlying physical condition [21]. Our finding that our rate of admissions from LTC to hospitals with organised stroke care was less than one third of stroke the incidence rate in LTC in Wales [6], which operates a similar model of healthcare and social care to Ireland is notable and of a perception that there is a discrepancy between the number of apparent stroke deaths identified by the audit and by the Central Statistics Office, vital statistics. This possible non-referral is compounded by the fact that, when admitted, these Long-Term Care residents are also significantly less likely to receive care in a stroke unit when we know that stroke unit care has been found associated with better outcomes in all sub-groups examined [8]. The substantially higher mortality found in our LTC referrals and controls not admitted to stroke unit my indicate either a poorer standard of care or a tendency to triage out patients with a perceived poorer predicted outcome prior to admission.

It is reassuring that admission from LTC was not itself associated with a worse outcome from stroke, suggesting that such admissions were not treated differently from those of people living in the community. It is also worth noting that the outcome of people admitted with stroke from LTC was not universally bleak with 30% of patients independent on admissions maintaining their independence on discharge. Although patients from LTC were less likely to be admitted to stroke units, there may have been other factors affecting this including need for isolation or palliative care. it is encouraging that these LTC admissions received the same level of intervention in terms of thrombolysis and swallow assessment as the controls form the community. There is increasing evidence that pre-morbid levels of disability should not in themselves necessarily preclude patients from intervention and should be judged from the perspective of prognosis and quality of life [23, 24].

In conclusion, whilst LTC residents represent <5% of admissions to hospitals with stroke services. Although this is a higher proportion than that of in the overall population ≥65 years living in LTC, it is lower than one might expect from published stroke incidence rates in LTC settings. More than three quarters of LTC residents suffering a stroke were discharged alive. In an era where options for treatment and intervention have increased, even those with background disability [23,24], care plans for LTC residents should be reviewed and include provision for appropriate transfer of patients to organised stroke care where appropriate treatment may reduce future impairment and disability.

## Data Availability

Data is available on application to the National Office for Clinical Audit.https://www.noca.ie/about-noca/access-to-audit-data/

## Statements and declarations

No author has any competing interests or funding in respect of this submission.

